# Knowledge, attitude, and practices of reporting adverse events following immunization among healthcare providers in Simiyu Region, Tanzania: A cross sectional study

**DOI:** 10.1101/2024.11.22.24317763

**Authors:** Raphael C. Kambona, Alphonce I. Marealle, Manase Kilonzi, Emmanuel Masunga, Peter Kunambi, Helfrid B. Ilomo, Ritah Mutagonda

## Abstract

**Introduction:** Immunization is the most cost effective health intervention in the history of mankind. Its role in reducing the morbidities and mortality due to vaccine-preventable diseases is significant. Despite vaccines being important in saving lives; adverse events following immunization (AEFI) do occur that need prompt detection, management, and then reported, analyzed, and the findings being shared with all stakeholders to ensure vaccine safety. The reporting of AEFI in Tanzania is still low. Health Care Providers (HCPs) are obliged to collect and report AEFI. This study assessed the knowledge, attitude, and practices of reporting AEFI among HCPs in Simiyu region, Tanzania.

**Methodology:** This cross-sectional study was conducted between May and June 2024. A total of 180 HCPs were enrolled, structured questionnaire was used to assess the knowledge, attitude, and practices of reporting AEFI among HCPs at primary health facilities. Statistical Package for Social Science (SPSS) was used for entry and data analysis, and the findings were summarized using frequency and percentages. Chi-square test was used to screen for factors associated with AEFI reporting while Poisson regression model was used to confirm the determinants.

**Results:** The median age of study participants was 32 (Inter Quartile Range; 29, 38) years. Among the HCPs, more than one quarter (28.89%) had adequate knowledge of reporting AEFI, and more than half (68.89%) had positive attitudes and good practices (62.22%) of reporting AEFI. Less than half of HCPs (48.3%) had encountered AEFI cases, and about two-thirds (69%) reported it.

**Conclusions and recommendations:** Despite the inadequacy of knowledge on reporting AEFI among HCPs, the attitude and practices of reporting AEFI among HCPs were promising. Based on the study findings, we recommend more training on vaccine surveillance, routine follow-up, and supportive supervision of AEFI surveillance for optimal AEFI reporting and vaccine safety surveillance in Tanzania.

## Introduction

Vaccination is one of the most successful public health interventions; globally it has significantly reduced morbidity and mortality associated with Vaccine Preventable Diseases (VPD). Worldwide 2 to 3 million deaths due to VPD are being prevented yearly(1–4). As the number of vaccines increased in routine immunization programs, the safety measures including Adverse Event Following Immunization (AEFI) reporting should also increase through passive surveillance of vaccines (3,5,6).

Even though vaccine production is closely monitored; each vaccine is accompanied by a kind of risk known as Adverse Event Following Immunization (AEFI). AEFI is any untoward medical event occurring following vaccination related to the vaccine administration and or its handling, and may not necessarily have a causal relationship with the vaccine use. The adverse event may be any unfavorable or unintended sign, an abnormal laboratory finding, symptoms, or disease (7,8).

Surveillance for AEFI can be active, passive, or stimulated. Active surveillance involves intensive follow-up of the cases, laboratory investigation, and questionnaires. In contrast, passive surveillance depends on individual, unsolicited reporting, the ability to identify, and the willingness of HCPs to report AEFI. Passive reporting of AEFI is the primary responsibility of HCPs for safety and post-marketing surveillance of vaccines (9–11). The main goal of AEFI surveillance is to prompt detect, manage, and take safety regulatory actions.

The reporting rate of AEFI in the WHO-African region is inconsistent and varies among member countries. The underreporting is attributed to weak AEFI monitoring systems, lack of guidelines and review committees, insufficient trained personnel, and inadequate stakeholder collaboration (12–15). In Tanzania, vaccine safety surveillance is managed by the Immunization and Vaccine Development (IVD) and the Tanzania Medicines and Medical Devices Authority (TMDA). However, even with this coordination, AEFI reporting remained suboptimal in the country (16).

This study assessed the knowledge, attitude, and practices of reporting AEFI among HCPs in Simiyu Region, it came up with the findings and proposed recommendations for improving AEFI reporting in Tanzania. The increase in AEFI reporting will increase vaccine safety within the country, promote immunization coverage, and reduce VPD.

## Methods

### Design and settings

This study used an analytical cross sectional design, it was conducted from May to June 2024 in primary health facilities providing vaccination services in Bariadi district in Simiyu Region. Bariadi district is located in the eastern lake zone of Tanzania; it comprises Bariadi District Council and Bariadi Town Council. The study involved Bariadi District Hospital, Bariadi Town Hospital, Songambele Hospital, and other 39 primary health facilities.

### Study population

This study involved Routine Immunization Service Providers, clinicians at outpatient department (OPD), and Pharmacy personnel working at primary health facilities in Bariadi district. A total of 221 Healthcare providers were included and signed the informed consent. Of 221 HCPs who consented to the study, only 180 (81.4%) responded fully to the questionnaire and took part in the final analysis.

### Data collection tools

Data were collected using a semi-structured questionnaire administered face-to-face to participants. The questionnaire was developed by adapting similar studies and modified to meet the study objectives (17,18). The questionnaire was pre-tested through the online survey and re-developed accordingly to fit data collection based on study objectives. The questionnaire consisted of 6 items on demographic information, 9 semi-structured questions on knowledge, 7 questions on attitude, and 6 questions on practices. Before conducting the interview; informed consent forms were provided to study participants.

### Data collection procedures

#### Administering the questionnaire

Study participants were recruited before conducting the study by the principal investigator and District Pharmacists. The district Pharmacist were working and supervising within their respective districts. Detailed information on the study was given to each participant, and raised questions were clarified before the participant was asked to sign the consent form. An average of 1 hour was given to participants to respond to questionnaire before collecting it back. After the consent, the questionnaire was given which was face to face administered and responses were provided accordingly.

### Data analysis

Both descriptive and inferential statistics were used to analyze the data collected, and continuous variables were presented as median and interquartile ranges. All categorical variables were presented in proportion and frequency. Level of knowledge, attitude, and practice were measured as the primary outcomes. The knowledge part consisted of 9 questions; the obtained proportion was multiplied by 100 to attain the percentage of knowledge for each participant. Knowledge was classified as “adequate” for scores ≥ 75%, and “inadequate” for scores less than 75% (19,20).

A set of 7 questions was used to ascertain the attitude of study participants, the responses to the question ranged from “strongly disagree, disagree, neutral, agree and strongly agree” in which the true statement was scored 1,2,3,4,5 concerning strongly disagree, disagree, neutral, agree and strongly agree statements. The false statement scored 5,4,3,2,1 marks for strongly disagree, disagree, neutral, agree, and strongly agree respectively. The proportion of attitudes for each participant was obtained by calculating the sum of all correct responses divided by all possible correct responses. HCP who scored marks between 21-35 was deemed to have a positive attitude on AEFI reporting, scored less than 21 marks was deemed to negative attitude on AEFI reporting (20).

There was a set of 6 questions that ascertained the practice of AEFI reporting, each question had 1 mark; the HCP who scored 3-6 questions was deemed to have good practice on AEFI reporting, scoring less than 3 questions was deemed to poor practice of reporting AEFI (20)(19). On Social demographic data, age was measured in years, duration of practice was measured in years 1-4 years was regarded as junior HCPs, 5-10 years was regarded as seniors, and more than 10 years was regarded as principals. Gender was measured by giving options such as male and female, professional and qualification based on the level of education ranging from certificate, diploma, and bachelor degree, and the level of the facility either Hospital, Health center, or dispensary. Chi-squared test of association and Poisson regression with robust standard errors model were used to observe significance between categorical variables in reporting of AEFI. Microsoft Excel and Statistical Package for Social Science (SPSS) version 23 were used for data entry and analysis. The confidence level was set at 95% and the p-value at 0.05. The data were cleaned and analyzed as per study objectives. All variables that were significant in the Chi-square were subjected to Poisson regression with robust standard error analysis to determine the relationship between demographic/other independent variables of the respondent on reporting of AEFI.

### Ethical consideration

Ethical clearance to conduct this study was obtained from the MUHAS-Institutional Review Board before conducting the study (Reference no. MUHAS-REC-03-2024-2066, and its approval amendment referenced DA.282/298/06/C/767)).Prior to commencement of the study, the participants were given information on voluntarily practice of participation, the confidentiality of the information provided were guaranteed, and freedom to withdraw in the study anytime whatever they feel were given without any penalties imposed. No participants names were indicated in the questionnaire, instead numbers were used to ensure confidentiality.

## Result**s**

A total of 221 Health Care Providers (HCPs) were recruited for this study, but 180 respondents completely filled out the questionnaire and were included in the final analysis which accounted Participant’s response rate of 81.4%.

### Socio-demographic characteristics of study participants

About half of the HCPs (56.7 %, n=102) were females. More than half (63.3%, n=114) were married, more than three quarters (78.3%, n=141) had college education, about half (53.9%, n=97) had a diploma as highest professional education, and more than one third (41.1%, n=74) were nurses. Participant’s median age was 32 years. Most of HCPs (67.8%, n=122) were middle-aged (≤35 years), more than one third (44.4%, n=80) had 1-4 years of working experience and most (76.1%, n=137) of HCPs were directly involved in routine immunization services. *(Table 1)*.

**Table 1.**
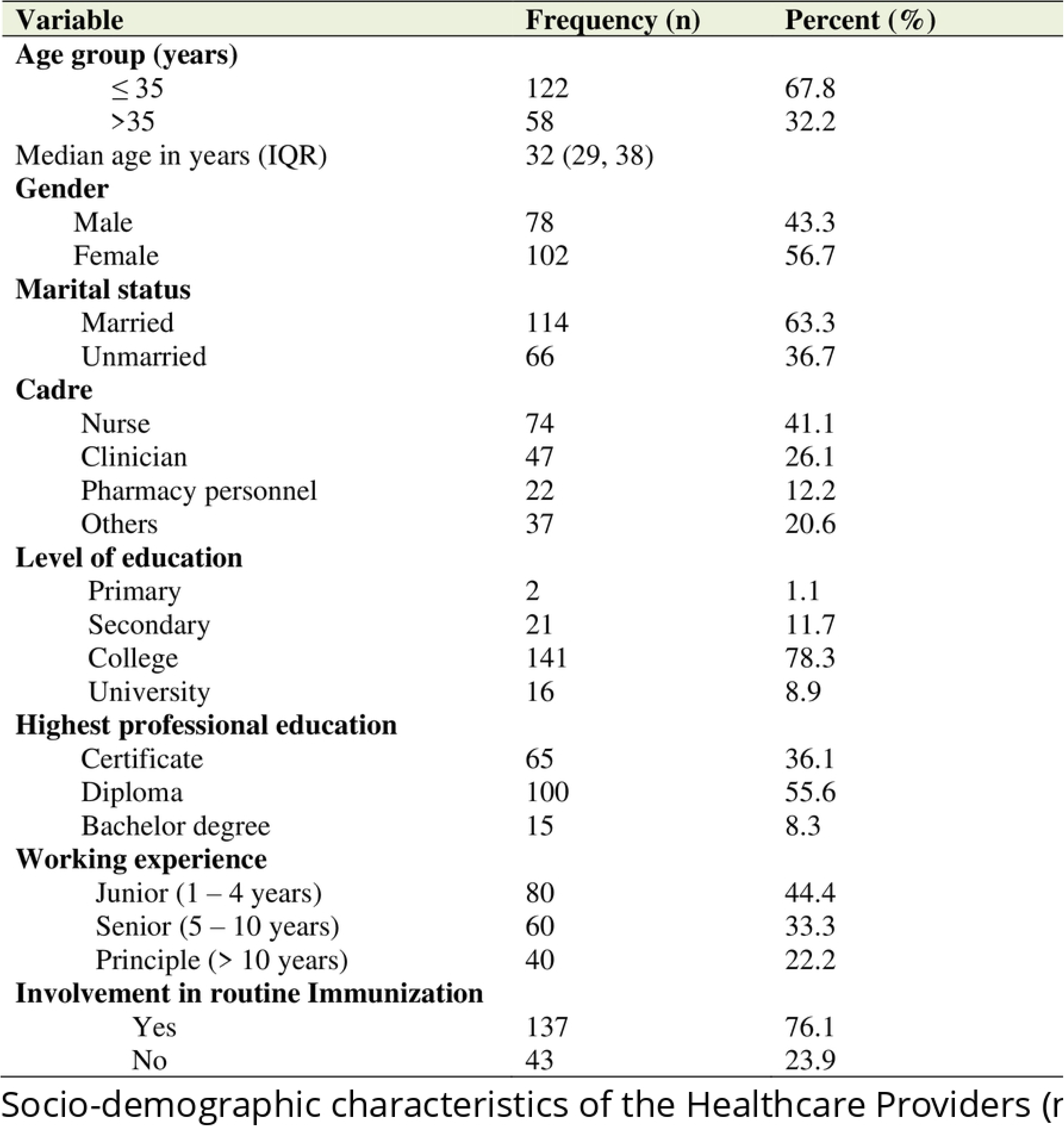

### Knowledge on AEFI reporting

The overall proportion of HCPs with adequate knowledge of reporting AEFI was 28.9% (n=52), while 71.1% (n=128) had inadequate knowledge. Most HCPs (72.8%, n=131) correctly defined AEFI. The majority of HCPs (95.6%, n=172) were aware of reportable AEFI cases which comprised serious and non-serious cases; only few participants (4.4%, n=8) considered only serious AEFI should be reported. Most of HCPs (92.8%, n=167) heard AEFI surveillance on reliable sources, and the majority (93.3%, n=168) heard about AEFI surveillance. Few participants (6.7%, n=12) knew various modalities of reporting AEFI, while more than three quarter of Participants (77%, n=139) used individual case safety report (yellow form) as the means of reporting AEFI. Few HCPs (9.4%, n=17) had the knowledge on the types of AEFI as classified by WHO based on vaccine product related, vaccine quality defect, immunization anxiety, immunization error, and coincidental. About one quarter of the participants (26.1%, n=47) knew AEFI cases subject for investigation, and most of the HCPs (90.6%, n=163) know AEFI can be prevented but among them only few HCPs (27.8%, n=50) know the methods to prevent AEFI. (*Fig 1)*

**Fig 1.**
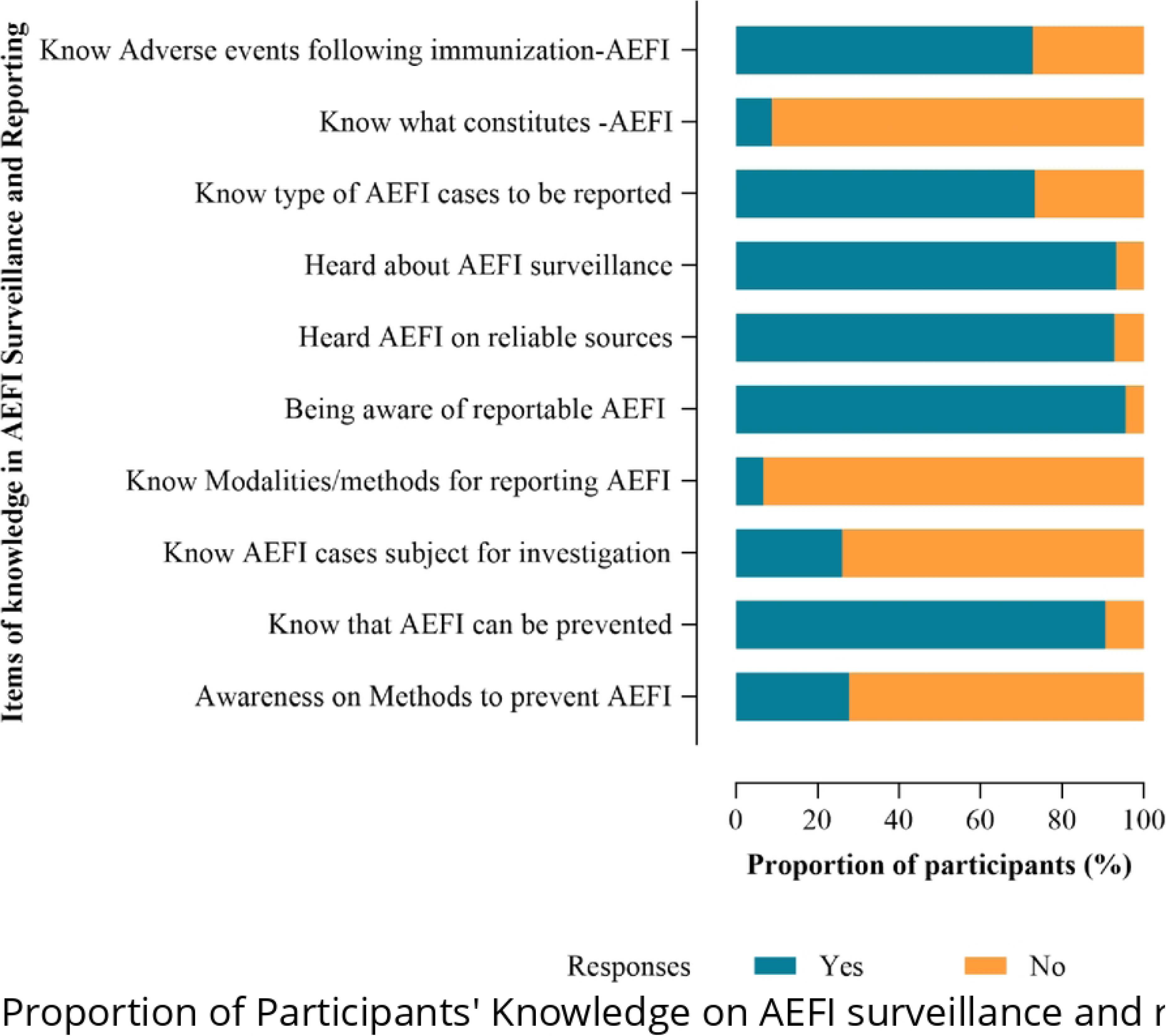

### Participant’s attitude on AEFI reporting

Of the 180 respondents, 96.7% (n=174) agreed on the importance of AEFI training towards improving AEFI reporting, and the majority of respondents (90.6%, n=163) agreed that it is their professional obligation to report AEFI. Most of respondents (96.7%, n=174) agreed that AEFI reporting improves patient care since prompt action and further management will be given to the Patient when AEFI is reported. Few respondents (18.3%, n=33) reported that AEFI creates additional work while others (13.3%, n=24) considered reporting AEFI as a time-consuming activity, and some (7.2%, n=13) could not report AEFI by fearing of being victimized by their bosses *(Fig 2).* Generally, the majority of participants (68.89%) had a positive attitude toward reporting AEFI.

**Fig 2.**
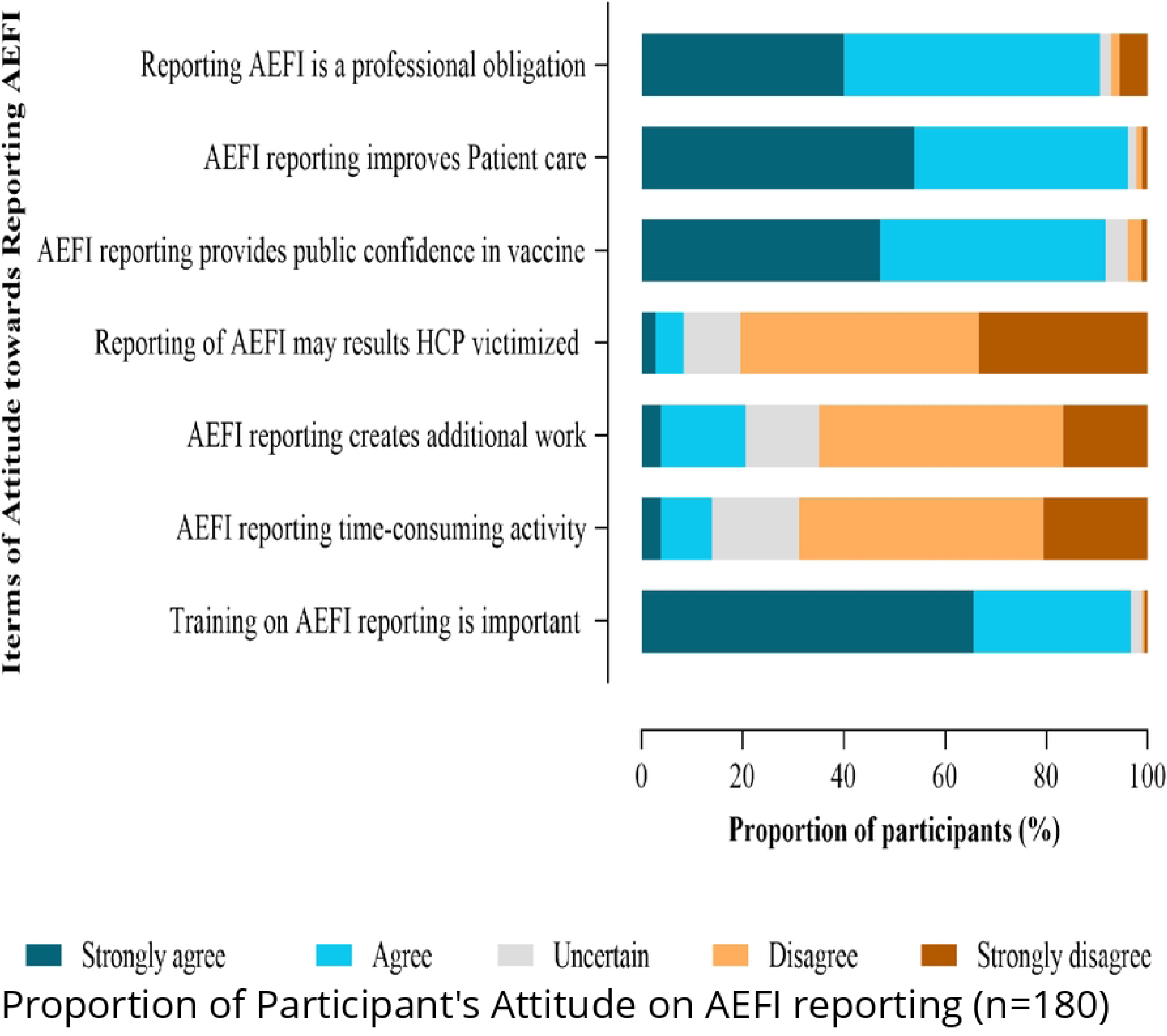

### Practices of reporting AEFI

The majority of the HCPs (62.22%, n=112) had good practices of reporting AEFI, and most (69.4%, n=125) elaborated the procedures of handling, managing, and reporting of AEFI. Less than half of HCPs (48.3%, n=87) encountered AEFI cases during their routine practices, and two third (69%, n=60) reported it. Less than half (47.2%, n=85) provided reasons for underreporting AEFI with the responses ranging from; not aware of the importance of reporting AEFI (43%,), don’t know how to report AEFI (3.5%), AEFI are self-limiting (30.4%), no follow up by the administrators (11.7%), fear of being victimized by their bosses (8.3%) and other reasons (3.0%). Most HCPs (96.7%, n=174) were unfamiliar with the variety of modalities of reporting AEFI. The mostly used method for reporting AEFI was individual case safety report (yellow form) (66.5%), other reporting modalities were phone calling to Tanzania Medicines and Medical Devices Authority (TMDA) /District (15.8%), using TMDA website (6.2%), Toll free number (5.3%), mobile application (3.8%), and email (2.4%). *(Fig 3)*

**Fig 3.**
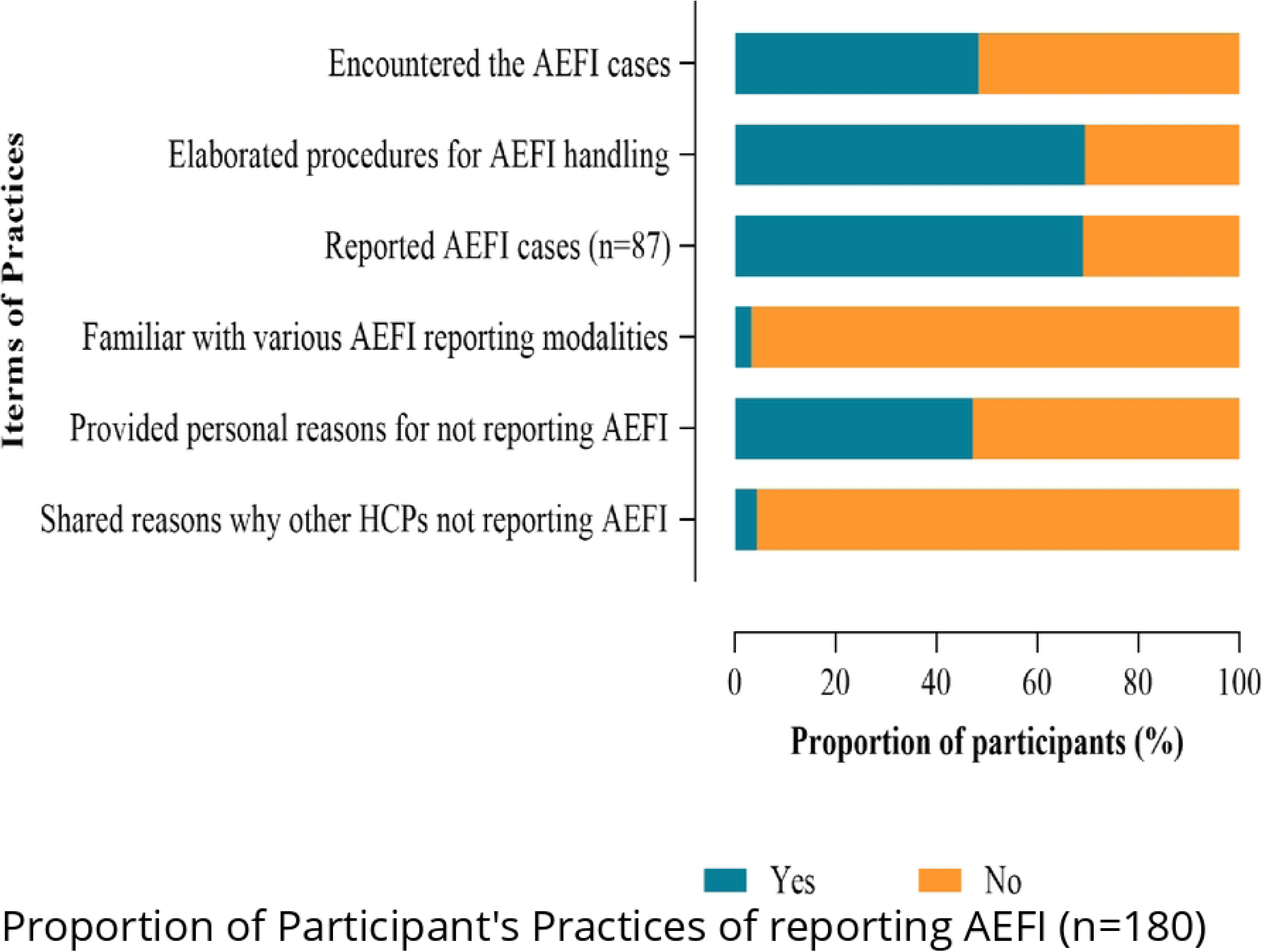

### Factors associated with Participants’ knowledge, attitude, and practices on reporting AEFI

Among the socio-demographic characteristics, Participants who reported AEFI did not differ from those who did not report AEFI with regard to age group, gender, marital status, cadre, involvement in routine immunization, and level of education. The highest professional education was statistical significantly associated with knowledge of reporting AEFI (p =0.002). (*Table 2*). Among the attitude characteristics, participants who reported AEFI did not differ from those who did not report AEFI with regards to age group, gender, marital status, cadre, level of education, working experience, involvement in Routine immunization services, and highest professional education. The knowledge was significantly associated with attitudes of reporting AEFI, the participants with knowledge of AEFI reporting were more likely to have positive attitude of reporting AEFI. (*Table 3*). Among the practice characteristics of participants, socio-demographic characteristics were not statistically significantly associated with the Practices of reporting AEFI. The practices of reporting AEFI were significantly associated with knowledge and attitude (p <0.05). Multivariable analysis showed participant’s age group ≤ 35 years (p=0.011), working experiences 1-4 years (p=0.025), working experience of 5-10 years (p=0.033), and adequate knowledge on reporting AEF (p <0.001) being significant associated with good practices of reporting AEFI *(Table 4)*.

**Table 2.**

Socio-demographic characteristics of the Healthcare Providers (n = 180)
| Variable | Knowledge on reporting AEFI |  | p – value |
| --- | --- | --- | --- |
|  | Adequate (%) | Inadequate (%) |  |
| **Age group (years)** |  |  |  |
| ≤ 35 | 34 (27.9) | 88 (72.1) | 0.661 |
| >35 | 18 (31.0) | 40 (69.0) |  |
| **Gender** |  |  |  |
| Male | 24 (30.8) | 54 (69.2) | 0.626 |
| Female | 28 (27.5) | 74 (72.5) |  |
| **Marital status** |  |  |  |
| Married | 29 (25.4) | 85 (74.6) | 0.180 |
| Unmarried | 23 (34.8) | 43 (65.2) |  |
| **Cadre** |  |  |  |
| Nurse | 18 (24.3) | 56 (75.7) | 0.401 |
| Clinician | 17 (36.2) | 30 (63.8) |  |
| Pharmacy personnel | 8 (36.4) | 14 (63.6) |  |
| Others | 28 (75.7) | 9 (24.3) |  |
| **Level of education** |  |  |  |
| Primary | 1 (50.0) | 1 (50.0) | 0.388 |
| Secondary | 5 (23.8) | 16 (76.2) |  |
| College | 39 (27.7) | 102 (72.3) |  |
| University | 7 (43.8) | 9 (56.3) |  |
| **Highest professional education** |  |  |  |
| Certificate | 9 (13.8) | 56 (86.2) | 0.004 |
| Diploma | 37 (37.0) | 63(63.0) |  |
| Bachelor degree | 6 (40.0) | 9 (60.0) |  |
| **Working experience** |  |  |  |
| Junior (1 – 4 years) | 24 (30.0) | 56 (70.0) | 0.093 |
| Senior (5 – 10 years) | 12 (20.0) | 48 (80.0) |  |
| Principle (> 10 years) | 16 (40.0) | 24 (60.0) |  |
| **Involvement in routine immunization** |  |  |  |
| Yes | 38 (27.7) | 99 (72.3) | 0.543 |
| No | 14 (32.6) | 29 (67.4) |  |

Factors associated with knowledge level on reporting AEFI

**Table 3.**
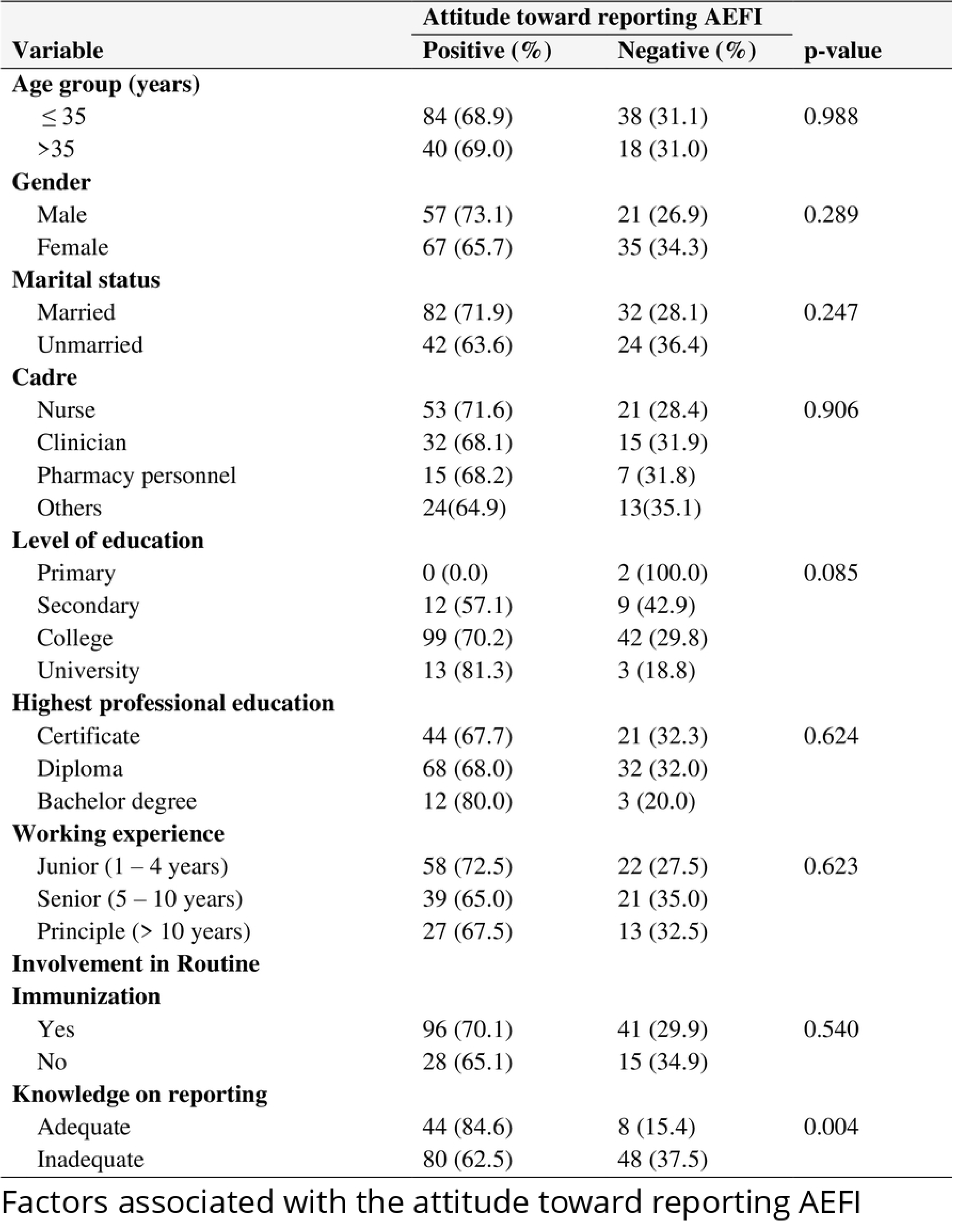

**Table 4.**
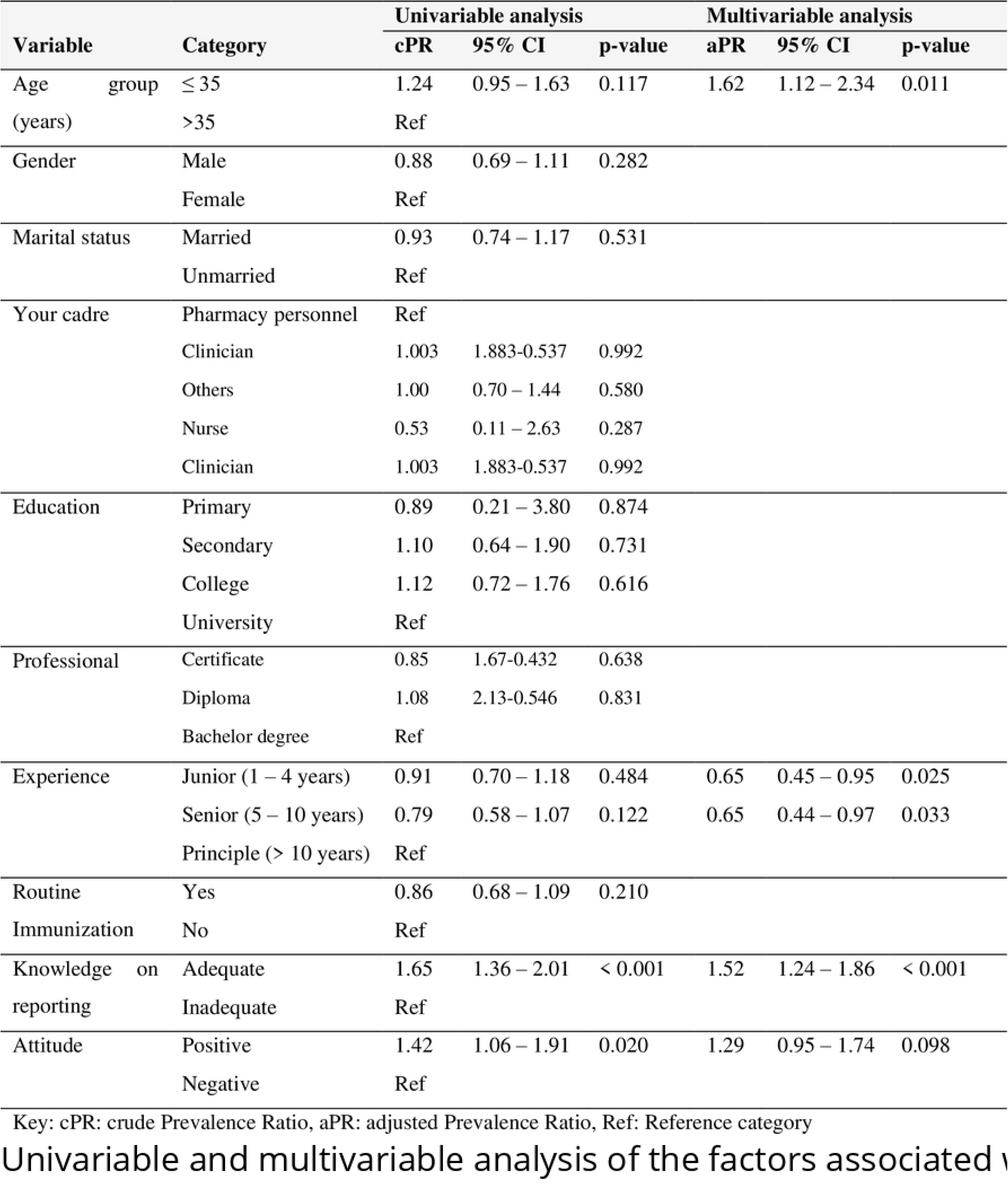

## Discussion

Vaccine safety surveillance is the cornerstone for vaccine safety and quality in immunization programs (1,19,20). Public mistrust of vaccines and vaccine hesitancy are the results of poorly detection and communication on AEFI; thus, Health Care Providers (HCPs) need to have good knowledge, attitude, and practices of reporting AEFI for building trust and optimal vaccine uptake by the community (21).Various studies on Health care provider’s knowledge, attitude, and practices of reporting AEFI have been conducted, but little is known in Tanzania (18,22,23). This analytical cross sectional study assessed knowledge, attitude, and practices of reporting AEFI among HCPs in Simiyu region, Tanzania.

In this study 28.89% (n=52) of HCPs were found to have adequate knowledge of reporting AEFI, which is similar to the same study conducted in Kenya by *Masika C. et al* in 2016 which showed 29.2% of HCPs had adequate knowledge. The finding of this study is contrary to the studies conducted in Nigeria and Ghana, in which 63.6% and 57.8% of HCPs had adequate knowledge on AEFI reporting respectively (19)((24). The inadequate of knowledge of reporting AEFI could be explained by inadequate supportive supervision on AEFI surveillance and the inadequate pre-service pharmacovigilance training in Tanzania (25).

In this study, the majority of HCPs (72.8%, n=131) correctly defined AEFI, this is similar to the same studies conducted in Ghana and in Nigeria in which 83.4% and 70% respectively correctly defined AEFI (24,26). This study shows HCPs characteristics of the highest professional education associated with good knowledge of reporting AEFI. Other demographic factors like age, marital status, involvement in routine immunization, working experiences, and cadres had no association with knowledge of reporting AEFI which is similar to the study by *Parella A* et (11).

Our study showed the majority of HCPs (68.89%, n=124) had a positive attitude towards reporting AEFI. The majority of HCPs (96.7%, n=174) agreed on the importance of training on AEFI reporting, this is similar to the same study conducted in Nigeria (19). The attitude of reporting AEFI was statistically significantly associated with the knowledge of reporting AEFI among HCPs (p=0.004) while other demographic characteristics like age, gender, marital status, and working experiences were not associated with the attitude of reporting AEFI. In our study, knowledge of AEFI reporting was significantly associated with the attitude of reporting AEFI (p=0.004), the finding is similar to the studies conducted in Ghana (2,24).

Some HCPs had a negative attitude and thought that; reporting AEFI creates additional work (18.3%, n=33), and reporting AEFI is a time-consuming activity (13.3%, n=24) as well as the fear of being victimized by their bosses (7.2%, n=13). The findings on negative attitudes replicate the study by *Malande.O et al* in Kenya, *Yamoah.P et al* in Ghana, *Laryea E. et al* in Ghana, and *Thomas.R et al* in India (2,13,21,24).

Surprisingly; working in routine immunization clinics was not associated with positive attitudes toward reporting AEFI, this is contrary to the similar studies in Ghana and Nigeria in which those HCPs working in Routine immunization clinics had a several-fold increase in good attitudes on reporting AEFI (20)(27). The reasons for working in routine immunization clinics having no association with positive attitudes on reporting AEFI could be influenced by myths during COVID 19 pandemic and political influences on vaccine safety (28). All stakeholders should ensure positive attitudes on reporting AEFI by HCPs for effective vaccine surveillance, since it is evident that; negative attitudes of HCPs do hinder AEFI reporting (4).

Our study shows that more than a half (62.22%, 112) of HCPs had good practices of reporting AEFI, the finding is similar to studies conducted in Nigeria (14,19).The majority (62.22%) of HCPs having a good practices of reporting AEFI could be attributed by previously sensitization meetings organized by Tanzania Medicines and Medical Devices Authority on reporting AEFI to HCPs. More than one-third (48.3%, n=87) of HCPs had encountered AEFI cases during their routine practices, of 87 HCPs who encountered AEFI, the majority (69%, n=60) reported it. The majority of the HCPs (69.4%, n=125) handled the AEFI cases appropriately upon encountering AEFI cases; they managed the case accordingly whether serious or non-serious and then reported it.

Less than a half of HCPs (47.2%, n=85) provided reasons for underreporting AEFI; of 230 provided reasons; the HCPs declared not aware of the importance of reporting AEFI (43%), AEFI are self-limiting (30.4%), no follow up by the administrators (11.7%), fear of being victimized (8.3%), they don’t know how to report AEFI (3.5%), and some provide other reasons (3.0%) which included; never encountered any AEFI case and it is not their responsibility to report AEFI. The findings on the reasons for underreporting AEFI reflect similar studies conducted in India, Zimbabwe, Ghana, Nigeria, and Kenya (13,15,19,21,24)

The provided reasons for the underreporting of AEFI should be worked on by the Government and all stakeholders to enhance AEFI reporting within the country for effective vaccine safety surveillance. This study shows more than three-quarter of HCPs (96.7%, n=174) did not use a variety of modalities for reporting AEFI which could had contributed to AEFI underreporting. Of modalities used for reporting AEFI; yellow form was mostly used (66.5%) compared to other modalities which include phone calling to National Regulatory Authority (15.8%), using website (6.2%), using toll-free number 080008110084 (5.3%), using mobile application (3.8%), and by using email (2.4%). The finding of this study is similar to the same study conducted in Nigeria in 2020 in which the majority of HCPs (95.1%) used Individual Case Safety Report (yellow form) as the main modality for AEFI reporting (19). Tanzania Medicines and Medical Devices Authority, and Immunization and Vaccine Development should ensure the utilization of other appropriate modalities for reporting AEFI including vigimobile for effective increase AEFI reporting and subsequent vaccine safety surveillance.

### Strengths and Limitations

The tool used for assessing participants’ knowledge, attitude, and practices was based on other colleagues’ and pharmacovigilance experts’ knowledge which justified its validity. This study faced several limitations including recall bias to HCPs when responding to some questionnaire. The principal investigator provided further information to verify the questions. This study was conducted only to HCPs and excluded the clients/parents, this is another limitation towards assessment of knowledge, attitude and practices of reporting AEFI since both HCPs and clients/parents are obliged to report AEFI. This gave one sided finding on AEFI reporting based on HCP’s perspective.

### Conclusions and recommendations

This study showed less than one third (28.9%, n=52) of HCPs had adequate knowledge of reporting AEFI. Despite the inadequacy of knowledge on AEFI reporting by majority of HCPs; the findings revealed more than a half HCPs (68.89%, n=124) had a positive attitudes and good practices (62.22%, n=112) of reporting AEFI. More than one third (48.3%, n=87) of HCPs had encountered AEFI cases during their routine practices, and majority of them (69%, n=60) reported it. The study findings revealed association between adequate knowledge of reporting AEFI with highest professional education, positive attitude of reporting AEFI with knowledge of reporting AEFI, and good practices of reporting AEFI with adequate knowledge. We do recommend the aforementioned reasons for underreporting, gaps on HCP’s knowledge, attitude, and practices to be intervened by relevant stakeholders for effective AEFI reporting and subsequent vaccine safety surveillance within the country.

### Supporting information

Questionnaire

Consent form

Ethical approval

## Data Availability

All data produced in the present study are available upon reasonable request to the authors

## Acknowledgment

We firmly acknowledge the real cooperation we received from Bariadi district executive director, Bariadi Town Director, District medical officers, District Pharmacists, Participants, and the entire staff of the School of Pharmacy, Department of Clinical Pharmacy and Pharmacology of Muhimbili University of Health and Allied Sciences.

## Author contributions

Conceptualization: Raphael C. Kambona

Data curation: Raphael C. Kambona

Formal analysis: Raphael C. Kambona, Peter Kunambi, Alphonce Marealle

Funding acquisition: Raphael C.Kambona

Methodology: Raphael C.Kambona, Manase Kilonzi, Emmanuel Masunga, Helfrid B.Ilomo

Supervision: Raphael C.Kambona, Ritah Mutagonda, Alphonce Marealle

Project administration: Raphael C.Kambona Validation: Raphael.C.Kambona

Writing-original draft: Raphael C.Kambona, Manase Kilonzi

Writing-review & editing: Raphael C. Kambona, Ritah Mutagonda, Alphonce Marealle, Manase

Kilonzi, Emmanuel Masunga.

